# Examining audiology students’ clinical reasoning skills with virtual audiology cases aided with no collaboration, live collaboration, and virtual collaboration

**DOI:** 10.1101/2020.07.21.20159426

**Authors:** Ramy Shaaban

## Abstract

The purpose of this study was to examine the difference in clinical reasoning skills of students when using virtual audiology cases aided with no collaboration, virtual collaboration, and live collaboration. The theoretical framework discussed five major approaches that could explain this effect; Situated Learning Theory, Social Development Theory, Scaffolding, Collaborative Learning, and Cognitive Load Theory. A quasi-experimental design was conducted at two U.S. universities to examine whether there is a significant difference in clinical reasoning skills between the three treatment groups using IUP Audiosim software. The clinical reasoning data were analyzed using One-Way ANOVA, and Tukey HSD for ANOVA. The results indicated that there was a significant difference in clinical reasoning skills between the three treatment groups. Further analyses are shown in the body of the study.

## INTRODUCTION

This study was designed to examine the clinical reasoning skills of students using electronic virtual patients aided with no collaboration, virtual collaboration, or live collaboration. While online simulations have been found to be essential in improving clinical reasoning skills, electronic virtual patients are still being evaluated and could be found to give improved results on the performance of students if aided with either virtual or live collaboration. This study attempted to add to the body of knowledge on this educational and training research topic in the field of medical education.

Electronic Virtual Patients (EVPs) are computer simulation programs that are essential components of medical education (Bateman, Allen Maggie, Kidd, Parsons, & Davies, 2012). EVPs are well known to enhance clinical reasoning skills in healthcare and medical education. Clinical reasoning refers to the process by which healthcare professionals collect cues and information to reach an accurate diagnosis of a medical case. Clinical reasoning, before the era of virtual patients, was enhanced by creating a collaborative learning environment where students could collaborate to discuss possible diagnoses, thus reaching the final diagnosis quickly and accurately.

The issue this study addressed was that the design of the current virtual patients used in medical training lacks collaborative reasoning, which is an important part of clinical reasoning. The study aimed to examine whether adding collaboration improves the performance of the students using the virtual patient and whether this effect differs when using virtual versus live collaboration. The following sections discussed the problem in detail, including research questions and hypotheses. A brief description of the study design also follows.

Although there is no solid definition for clinical reasoning, Barrows and Feltovitch (1987) described clinical reasoning as a problem-solving process used by doctors. Clinical reasoning is also described as cognitive skills that a physician or a health professional needs to master to diagnose a clinical case (Custers, 2018). Due to the ambiguity of this term in the literature, Simmons (2010) conducted a concept analysis aiming to describe the term clinical reasoning. She defined it as a “complex process that uses cognition, metacognition, and disciplineJspecific knowledge to gather and analyze patient information, evaluate its significance, and weigh alternative actions” (Simmons, 2010).

Since there are policies that prevent direct contact with patients without a medical license, educators have developed alternative ways to overcome this problem. One of the most used tools is simulation-based education. Medical educators used to call these simulation-based tools virtual patients. The most unique and cost-effective function of virtual patients is to facilitate and assess the development of clinical reasoning (Cook & Triola, 2009). Cook and Triola (2009) concluded that virtual patients should be designed to enhance clinical reasoning skills; thus, more research is needed to inform how to use virtual patients effectively.

Virtual patients provide predefined scenarios for clinical cases where students can engage in problem-solving activities to find a final and most accurate diagnosis (Ellaway, Candler, Greene, & Smothers, 2006). These problem-solving activities are known to enhance the students’ clinical reasoning skills or their ability to diagnose a medical case (Boshuizen et al., 2018). Thus, virtual patients could improve medical student performance by the problem-solving approach they mainly use in the clinical learning process.

Research on clinical reasoning started in the 1970s, taking a psychological perspective, and exploring how clinicians think when dealing with clinical cases (Pelaccia, Tardif, Triby, & Charlin, 2011). However, a few pieces of literature addressed how to improve clinical reasoning through instructional design (Braun et al., 2019). Rencic, Trowbridge, Fagan, Szauter, and Durning (2017) conducted a study to describe the clinical reasoning curricula at US medical schools. They found that most of the medical students enter the clerkship or the clinical training with only poor to fair knowledge of key clinical reasoning concepts (Rencic et al., 2017). Moreover, most medical institutions do not provide specific education for clinical reasoning (Rencic et al., 2017). Lastly, the lack of time available, as well as the lack of expertise in teaching clinical reasoning, are identified as barriers for medical institutions to provide clinical reasoning training (Rencic et al., 2017).

On the other hand, about 75% of diagnostic errors in medicine can be attributed to the physician’s cognitive failure (Norman et al., 2017). Cognitive psychologists tried to understand the cause of clinical errors by analyzing the clinical reasoning process by studying cognition (Graber, 2005). In 1974, Evans suggested a theory to examine the clinical reasoning process based on cognition, called the Dual Process Model (Wason & Evans, 1974). According to the model, human thinking has two distinct types of processes; heuristic processes, and analytic processes (Wason & Evans, 1974). The heuristic process is the first step of clinical thinking where an individual filters relevant information from a current situation (Wason & Evans, 1974). The second process takes this relevant information and uses it to judge the clinical situation (Wason & Evans, 1974).

In 2017, Norman et al. (2017) identified the cause of errors in clinical reasoning among healthcare professionals and aimed to determine whether the error came from the system one heuristic process or system two analytical process of thinking. The study found that healthcare professionals are vulnerable to commit mistakes in diagnosing cases due to three factors: cognitive bias, knowledge deficits, and dual-process thinking (Norman et al., 2017). In their study, they identified strategies to reduce the general error; one of these strategies is to use Kahneman’s (1963) model to “slow down and ask for reinforcement from system one.” This strategy led to significantly reduce cognitive bias in clinical reasoning (Norman et al., 2017).

While many pieces of literature address clinical reasoning on an individual basis, some literature examined the role of collaborative reasoning and claimed collaborative reasoning is qualitatively superior to individual reasoning (Geil, 1998). With the adoption of a student-centered learning paradigm, educators started to implement collaborative tools into electronic education and distant learning (Geil, 1998). The use of technology in collaborative learning needs a model of the nature of the anticipated collaboration (O’Donnell & O’Kelly, 1994). Existing literature described and analyzed the effect of collaborative learning in many ways, but many theoretical approaches were not successful in addressing exactly how peer learning can affect the learning process (O’Donnell & O’Kelly, 1994).

## Research Questions and Hypotheses

The purpose of the study was to examine whether collaborative learning has a positive effect on clinical reasoning skills.

- RQ1: Are there significant differences in terms of clinical reasoning skills between students using virtual patients aided with no collaboration, virtual collaboration, and live collaboration?
  - H0-1: There are no significant differences in terms of clinical reasoning skills between students using virtual patients aided with no collaboration, virtual collaboration, and live collaboration.

## Research Design

This study used a quasi-experimental design method to examine the clinical reasoning skills of the subjects. The subjects in this study were audiology students at a public university in Western Pennsylvania and another public university in Kansas. The experiment aimed to examine the differences in performance when the subjects use virtual patients with no collaboration, virtual collaboration, and live collaboration.

The experiment included three groups: the first group was the control group, which used virtual patient cases with no collaboration. Members of this group were not allowed to talk to each other while solving the cases. The second group received the same virtual patient cases but can collaboratively solve the cases through live classroom discussions. The third group received virtual patient cases with an online communication tool that enables them to collaborate virtually in solving the cases. The clinical reasoning skills were measured by analyzing the scores resulting from the virtual cases. These computer simulations can grade students and provide an overall score on their performance.

Students of communications disorders departments at both universities were the subjects of this study. Virtual cases given to them were audiology cases provided by the department. The sample was a convenience sample from audiology students taking audiology classes in the communications disorders departments.

## Significance of the Study

This study aimed to shed light on the need to improve the design of current virtual patients to be more effective in reducing clinical errors and enhancing clinical reasoning skills. The assumption was that promoting collaborative clinical reasoning rather than individual reasoning can overcome the current problems. Moreover, the study highlighted the difference in the efficacy between virtual and live collaboration tools embedded in the design of virtual patients.

This study aimed to examine the effectiveness of virtual collaborative reasoning in comparison to live collaborative reasoning, which gave an insight into the efficacy of virtual collaboration, thus recommending design modifications for new virtual patients. The study proposed a new design concept to combine collaboration with scaffolding aiming to enhance the efficacy of virtual patients in improving students’ clinical reasoning skills.

## Limitations of the Study

The study examined the efficacy of collaborative reasoning on audiology students in the communications disorders departments at two universities. Since audiology was the target specialty, this study cannot be generalized to cover other specialties. However, this study could be replicated in future research to examine the effect of collaboration on clinical reasoning skills in other specialties.

On the other hand, few studies examined the role of collaborative reasoning in overcoming clinical errors. Future studies are needed to investigate the significance of virtual collaboration in reducing clinical errors. Virtual collaboration embedded in virtual patient simulations should be examined for its efficacy in enhancing student clinical reasoning skills.

Another limitation was that the virtual collaboration tools that were used in this study were in the form of an online chatroom and file-sharing tools embedded in the virtual patient software. There are many forms of virtual collaboration tools such as discussion boards, forums, and video conferencing. Future studies could examine other forms of online virtual collaboration.

This study was done with audiology students in two universities, and also the experiment was repeated multiple times. This study could be replicated to examine audiology students in other schools that have different characteristics from the examined ones.

## Theoretical Framework

After the emergence of the constructivist view of learning and instructional design, constructing knowledge via active learning became prominent throughout the past 30 years (Streveler & Menekse, 2017). Constructivist learning is described as the active process where the learner creates the meaning from experience instead of just acquiring it (Ertmer & Newby, 2013). The constructivist theory claims that people build their analysis of the surrounding world from the knowledge they acquire in a specific context (Ertmer & Newby, 2013). With innovations, we see it is now imperative to stress that any innovation in technology must also be integrated with innovation in pedagogy (Colbert & Chokshi, 2014).

### Situated Learning and Social Development Theory

Situated learning is a human-centered approach that puts social context into the learning process. Through situated learning (SL), learners construct their knowledge through embedded scenarios, situations, and contexts (Anderson, Reder, & Simon, 1996). The Situated Learning Theory states that “every idea and human action is a generalization, adapted to the ongoing environment; it is founded on the belief that what people learn, see, and do is situated in their role as a member of a community” (Lave & Wenger, 1991).

The learning in the SL approach is a social process that can be constructed by the interactions among learners (Anderson et al., 1996). Linking the knowledge that needs an acquisition to the practice is the key to the situated learning approach (Lave, 2009). Without this link, the problem of inert knowledge can appear (Enkenberg, 2001). Inert knowledge is the acquired knowledge that learners have but cannot use because there is no definitive process for recalling it in the practical field (Whitehead, 1929).

Situated learning changes the role of the learner from a passive observer to an active participant in the learning process (Anderson et al., 1996). Being an active member in the learning process, enables learners to be more engaged in this process allowing them to express their ideas and use their cultural characteristics to enhance the learning experience (Johnson & Johnson, 2008). With the above concept, situated learning serves as an ideal approach for learner-centered education.

When mentioning situated learning theory, it is important to mention Vygotsky’s Social Development Theory (Vygotsky, 1980). In his theory, Vygotsky states that cognition and knowledge acquisition develop primarily from communication and social interaction (Jaramillo, 1996). Social interactions play a fundamental role in developing the cognitive abilities of the person (Jaramillo, 1996). There are two steps Vygotsky mentioned in the process of social-based cognitive development; the inter-psychological level where the person acquires knowledge through interacting with people, and intra-psychological where the person is cognitively self-developed from the inside (Manning & Payne, 1993).

To align Vygotsky levels of cognitive development with the theory of situated learning, learners go through two steps to acquire the ultimate knowledge and practice: the first level where the learner engages in active learning with other learners, and the second level where the learner develops his/her interpretation of the knowledge from inside him/herself (Andrade, Jaques, Vicari, Bordini, & Jung, 2001). This constructivist way of thinking about the learning process changes our view towards how knowledge is best transferred. In this situation, learning is a process of self-understanding, self-contextualizing knowledge based on the current experience. Thus, it is vital to shift our interest from a teacher-centered approach in the learning process to the learner-centered one. Moving from teacher-centered to learner-centered process changes the role of the teacher from being the source of information and directing personnel to become a facilitator and the support person.

On the other hand, instructional designers under this approach design learning environments that enable learners to construct knowledge based on social interactions and self-experiences (Mattar, 2018). Constructing knowledge through social interaction can be in the form of designing discussion sessions between learners, designing collaborative activities that enable learners to engage and reflect on the learning process (Mattar, 2018). Problem-based learning and case-based learning are other forms of situated learning approaches (Mattar, 2018).

Another term that is part of the situated learning approach is the community of practice. The community of practice is the social network of learners that are engaged in the learning process and work collaboratively to acquire the desired skill (Cruess et al., 2018). The community of practices provides situated context for their members to interact and negotiate with implicit and explicit meanings (Cruess et al., 2018).

### Computer-Based Situated Learning Approach

Situated learning nowadays can be implemented in the learning process via computer tools (Pérez-Sanagustín, Muñoz-Merino, Alario-Hoyos, Soldani, & Kloos, 2015). It is also common that instructors and course designers try to produce interactive computer media, allowing users to interact with computers the same way they interact with others in real life (Pérez-Sanagustín et al., 2015). Computer tutors, games, and interactive web applications are common forms of computer tools that use the situated learning concept (Pérez-Sanagustín et al., 2015).

Situated learning is becoming more involved with technology, given the fact that technology became very accessible to everyone (Mattar, 2018). In Computer-based interactive applications, the interaction happens through programmed, scripted computer interactions instead of human interactions, which is known as “Human-Computer Interaction” (Kim, 2015). Simulations enable learners to immediately apply what they have learned in a simulated real-life experience (McGuire & Babbott, 1967). Simulations can even provide a more effective situated learning experience than the traditional human-based ones, as simulations can allow learners to interact with broader and more distant communities, which cannot happen in the traditional situated learning setting (Jonassen, Davidson, Collins, Campbell, & Haag, 1995).

Computer applications, in their standard forms, are linear models that allow learners to acquire knowledge in a direct additive way (Halavais, 2016). However, using the natural learning process, the chaotic nature of it becomes clear (Halavais, 2016). We cannot claim that natural learning is entirely chaotic, but rather is a form of “organized chaos” (Somerville & Green, 2011). In real learning, people learn through physical interactions and perceptions of the surrounding world, without considering specific linear form to acquire the knowledge. The situated learning approach emphasizes this process of changing the learning experience from an additive linear model to a more real learning model (Young, 1995). So, how can this approach be applied in the computer-assisted learning experience?

Using situated learning as a human-centered model is essential in converting the learning experience from teacher-centered to student-centered one. Embedding situated learning approaches in online educational applications is very important for creating a more real life-like learning experience (Mattar, 2018). Designers use multiple tools to implement this approach in the online learning experience (Pérez-Sanagustín et al., 2015). Collaborative learning is one of the common forms used as an application of the situated learning approach (Pérez-Sanagustín et al., 2015). In online collaborative learning, learners use computer-based online tools to interact and collaborate with others (Pérez-Sanagustín et al., 2015). Thus, not only situated learning is vital in the traditional settings of the learning process, but it is also essential in the simulation and virtual learning experience.

Collaborative computer-based tools are numerous such as chatrooms, file sharing tools, discussion boards, blogs, social media tools, and online forums (Halavais, 2016). Embedding these tools into online learning activities can change the learning process from being a linear, additive process into a more realistic process (Chang et al., 2017). When learners interact via online tools, they can build-up knowledge that is based on their experiences and perceptions of the surrounding environment, thus building-up the desired constructivist approach (Pérez-Sanagustín et al., 2015).

In addition to collaborative learning, which makes the technology a tool for communication, rather than an educational tool, scaffolding is used to complete the situated learning experience (Lin, Hou, & Chang, 2020). Scaffolding is the assistive process given to the student throughout the learning experience (Braun et al., 2019). Embedding scaffolding into online learning can guide the student to the correct learning pathway (Braun et al., 2019).

The most common computer tools used to embed scaffolding into online learning is designing pre-scripted, interactive activities in the form of problem-solving sessions (Gonulal & Loewen, 2018). Building up applications that engage learners in an interactive session with customized feedback prompts based on learner’s interactions is the ultimate implementation of scaffolding in the online learning experience (Lin et al., 2020). In the following paragraphs, collaborative learning and scaffolding are discussed in depth. The effect of scaffolding and virtual collaboration on students’ critical thinking and clinical reasoning skills is also explored.

### Collaborative Learning and Scaffolding

In collaborative learning, learners are the center of the learning process as learners act as bricks that form the collaborative learning process. In their study about medical education, Colbert and Chokshi (2014) highlighted the importance of collaborative learning in that “As an educational tool, we believe that technology must be utilized in a manner that helps medical students develop into high-functioning communicators, collaborators, problem-solvers, and team players.” (*p*. *1584)* Collaborative learning is the learning process that happens when peer students collaborate to acquire knowledge (Greenwald et al., 2017).

Collaborative learning enhances the reasoning skills of the students by reducing clinical errors (Braun et al., 2019). Seventy-five percent of physicians’ clinical errors come from cognitive failures (Norman et al., 2017). Thus, collaborative learning could be a suitable solution to reduce clinical errors by enhancing clinical reasoning.

Going back to scaffolding, using scaffolding along with collaborative learning can have a potential solution for the challenges of acquiring clinical reasoning skills. Scaffolding is one of the conventional approaches used to improve the clinical reasoning process (Benson, 1997). Braun et al. (2019) mentioned multiple forms of scaffolds used to improve clinical reasoning: representation prompt where students imagine the hospital setting and write how they can present the clinical case to their colleagues, and structured reflections where the students are asked to reflect on their most-likely diagnoses (Braun et al., 2019). Thus, scaffolding is an assistive technique that can accompany students during the learning process to provide them with directions during acquiring clinical reasoning skills.

Scaffolding, on the other hand, is a process of guiding and assisting students throughout the learning process until reaching the goals. There are multiple types of scaffolding. The main two types are hard scaffolds and soft scaffolds (Shin, Brush, & Glazewski, 2017). Hard scaffolds are the pre-scripted assistive tools (Shin et al., 2017). During the online learning process, hard scaffolds can be in the form of custom feedback prompts that appear based on student’s inputs (Braun et al., 2019). Hard scaffolds aim to guide the student throughout the learning process to ensure keeping the student on the right track (Braun et al., 2019).

Soft scaffolds are another primary type of scaffolding. Soft scaffolds are dynamic, adaptive, assistive tools that are given to the student throughout the learning process. Soft scaffolds can be in the form of collaborative tools embedded in the online learning experience (Shin et al., 2017). Collaborative tools such as chatrooms can guide students throughout the learning process and can also keep them on track, but unlike hard scaffolds, they are adaptive; they can change according to student’s performance (Braun et al., 2019).

By virtual collaboration, researchers are referring to the tools that can be embedded in the online, clinical, simulation experience. An example of a virtual collaboration tool is the online chatroom (Cai et al., 2017). Chatrooms enable students to collaborate to solve challenging clinical cases (Leng & Gijlers, 2015). Creating a collaborative learning environment during the online simulation process can lead to enhancing clinical reasoning skills (Halavais, 2016).

### Cognitive Load Theory

In 1988, John Sweller developed Cognitive Load Theory based on the widely accepted model of human information processing that was developed by Richard Atkinson and Richard Shiffrin in 1968. The theory assumes that there are two types of knowledge; the biologically primary knowledge, which is the knowledge gained by an individual naturally without having to be acquired, and the secondary knowledge is the knowledge that can be acquired through instruction (Kirschner, Sweller, Kirschner, & Zambrano R., 2018). When addressing secondary knowledge, human cognition requires ample space to store information (Sweller, 2011).

Kirschner et al. (2018) argued that instructional design could be used to reduce the cognitive load. Before Sweller, and back in the 1950s, G. A. Miller was the first to research the field of cognitive science and suggests that working memory has a limited capacity (Sweller, 2011). There are three types of cognitive load: intrinsic cognitive load, germane cognitive load, and extraneous cognitive load (Chandler & Sweller, 1991). Intrinsic cognitive load is the inherent level of difficulty that results from a specific instructional topic. Extraneous cognitive load is generated as a result of specific instructional design. Germane cognitive load is defined as the working memory resources dedicated to dealing with intrinsic cognitive load (Sweller, 2011). Kirschner et al. (2018) argued that increasing the amount of interactivity leads to increase intrinsic cognitive load and extrinsic cognitive load.

On the other hand, collaborative learning occurs when two or more students participate in a shared learning process that requires interaction between those students (Kirschner et al., 2018). This shared learning process is usually provided and administered by the instructor. Two theoretical frameworks explain how collaborative learning affect cognitive load by reducing the interactivity; mutual cognitive interdependence and group cognition (Kirschner et al., 2018).

The mutual cognitive interdependence principle suggests that collaborative learning creates what is called a “collective working memory,” which develops through communicating between the interacting group, which leads to mutually shared cognition (Wegner, Giuliano, & Hertel, 1985). In addition to the cognitive interdependence, the theory of group cognition, which suggests that when a group of students participates in a cognitive process, the unit of analysis resulted from the group cognition becomes more substantial than the individual mind, so a group of working memories would process any interacting elements instead of single working memory (Stahl, 2005). Collaboration in this situation becomes a scaffold, thus reduces the extraneous cognitive load (Kirschner et al., 2018). This concept explains how collaborative learning can reduce the load, thus enhance clinical reasoning skills.

This chapter provided a review of the foundational as well as the current research in the field of collaborative reasoning, especially, the clinical reasoning. Scaffolding and collaborative learning are essential to clinical reasoning skills. The theoretical framework in this chapter discusses how clinical reasoning skills could be enhanced through scaffolding and collaborative learning. Theories such as cognitive load theory, situated learning, and mutual interdependence in this chapter were combined to explain the design model introduced in this study, which embeds collaborative and scaffolding tools into the virtual learning process to enhance students’ clinical reasoning skills.

## Research Design

The study used a quasi-experimental design method to examine the clinical reasoning skills of subjects. The experiment aimed to examine the significance of the differences in terms of subjects’ performance regarding their clinical reasoning ability to solve virtual clinical cases. Before running the experiment, a simulation program was developed and customized to have virtual collaboration tools embedded in it.

The quasi-experimental design was used to examine the clinical reasoning skills. Subjects must take a simulation session and solve virtual clinical cases in a computer lab, and their grade should be recorded and used as data that represents the level of clinical reasoning.

The following sections talk about each step in detail. First, the simulation program was reviewed in detail, and then the experiment was discussed with details about the experiment settings, then the sampling and recruitment were discussed. The following section provides details about how the study was conducted.

### Audiology Simulation Program

For this study, a simulation program was developed using Adobe Animate as the leading software to design the interactions, Google Forms as a tool for scaffolding and problem-based activity, and WordPress as a web host for the simulation session. The simulation was online and in a 2D format. It was formed of an interactive simulation for the clinical tools used to diagnose the clinical cases, and interactive problem-solving activity in the form of branched questions with scenarios that change according to the choices chosen by the student, and a grading system to record the students’ grades and save them in an excel sheet.

For this experiment, two virtual cases were developed. The cases had identical steps and activities but were different in the scenario and the case design. Because students are different and have different knowledge and experience using computers and web tools, the first case was straight forward to enable students to know how to use the simulation and to bring all the students to the same point when dealing with the second case. The second case was more challenging, aiming to test the students’ clinical reasoning skills.

The two modules designed for this experiment had the same process, the first module was a virtual case of a patient with an easy diagnosis. Easy diagnosis in this situation means the student had the needed clues and led to an accurate diagnosis with just simple reasoning. The case starts with mentioning the name, the age, the occupation, and the medical history of the patient. From this information, the student should get some clue about a wide range of diagnoses. The case then provided questions to the student to provide continuous assistance throughout the process. This is considered the hard scaffolding part of the case. The examination/investigation part of the module consisted of test results that were given to the student directly, the student just needs to look at these results and relate it with the previous case information to reach a diagnosis.

The second module had the same process. The difference was in the complexity of the case. In this module, there were some indirect clues, the student should think carefully to reach the correct diagnosis. It started with the same page of case information. In this case, the case information had hidden clues students should pay attention to them to get the correct diagnosis. In the experiment/investigation part, instead of giving the student the test results directly as the first module, the student enters an interactive simulation session, where the student gets a simulation of the clinical devices used in the audiology clinic such as the audiometer simulator (figure 8). The student used this interactive session to get the result themselves instead of getting the results directly.

Each case started with a history-taking session (See Figures 5, 6, and 7), which consisted of a problem-solving multiple-choice set of questions that enabled the student to obtain a patient’s history and think about the possible differential diagnoses. During the problem-solving session, hard scaffolds were embedded in the session in the form of scripted feedback. When the student gets the answer wrong, a popup message appears, letting him/her know the answer is wrong, giving them a reason why this answer is wrong, and asking them to try again — designing these hard scaffolds aimed to keep the student on track until they reach the final diagnosis. However, any wrong answer the student chose was taken into consideration in the overall grade.

**Figure 1.**
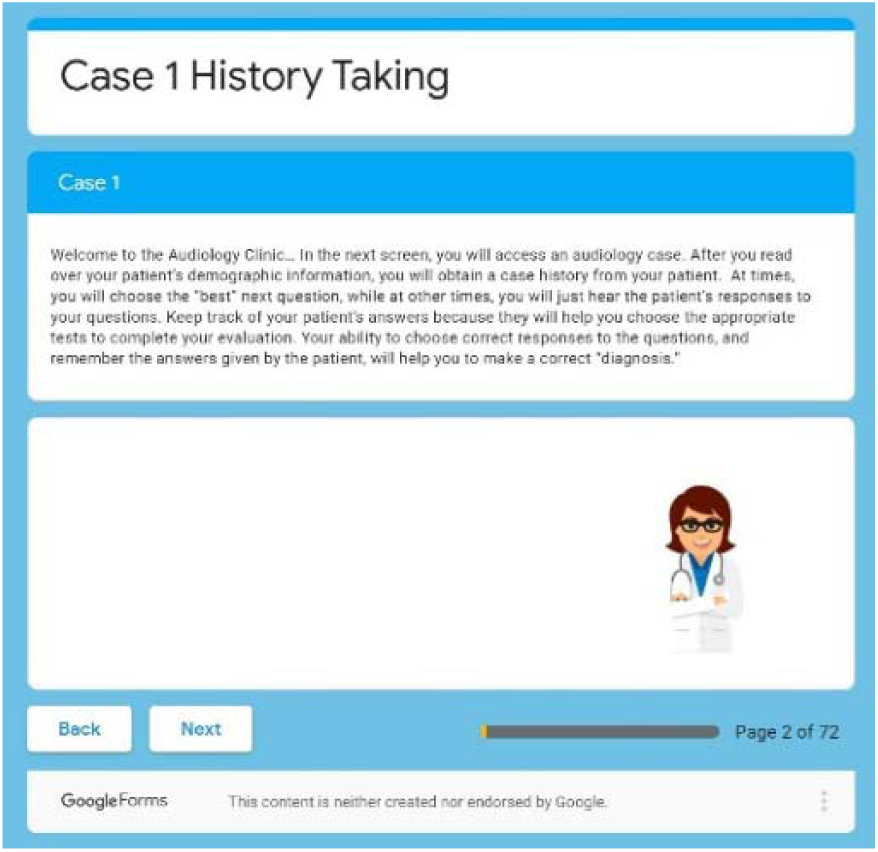
History taking section - First screen.

**Figure 2.**
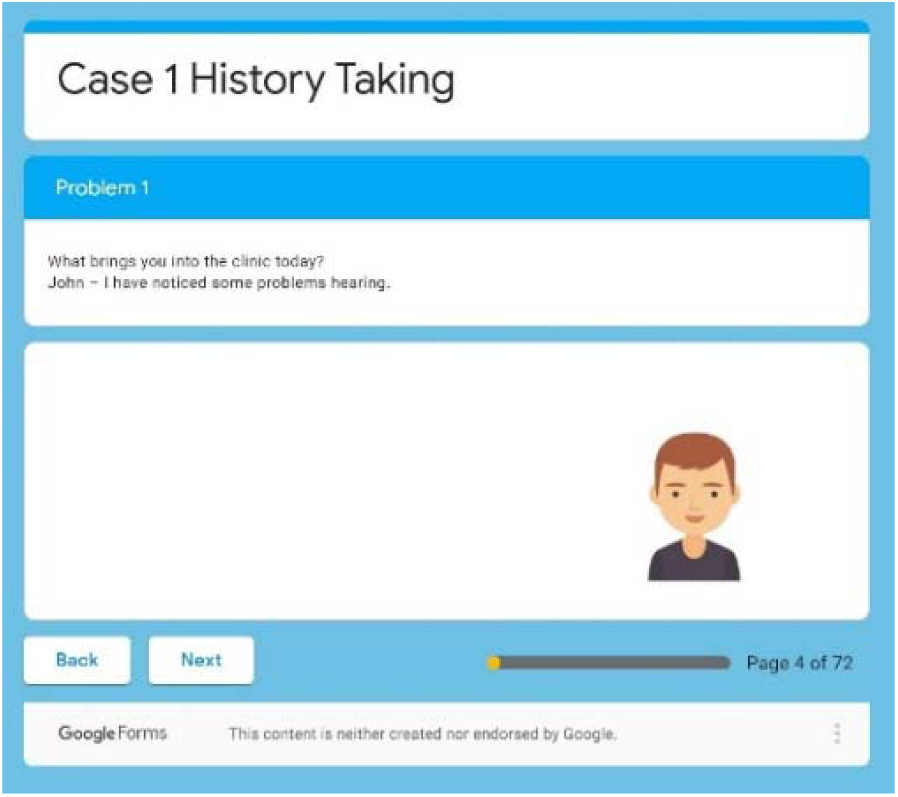
History taking section - Second screen.

**Figure 3.**
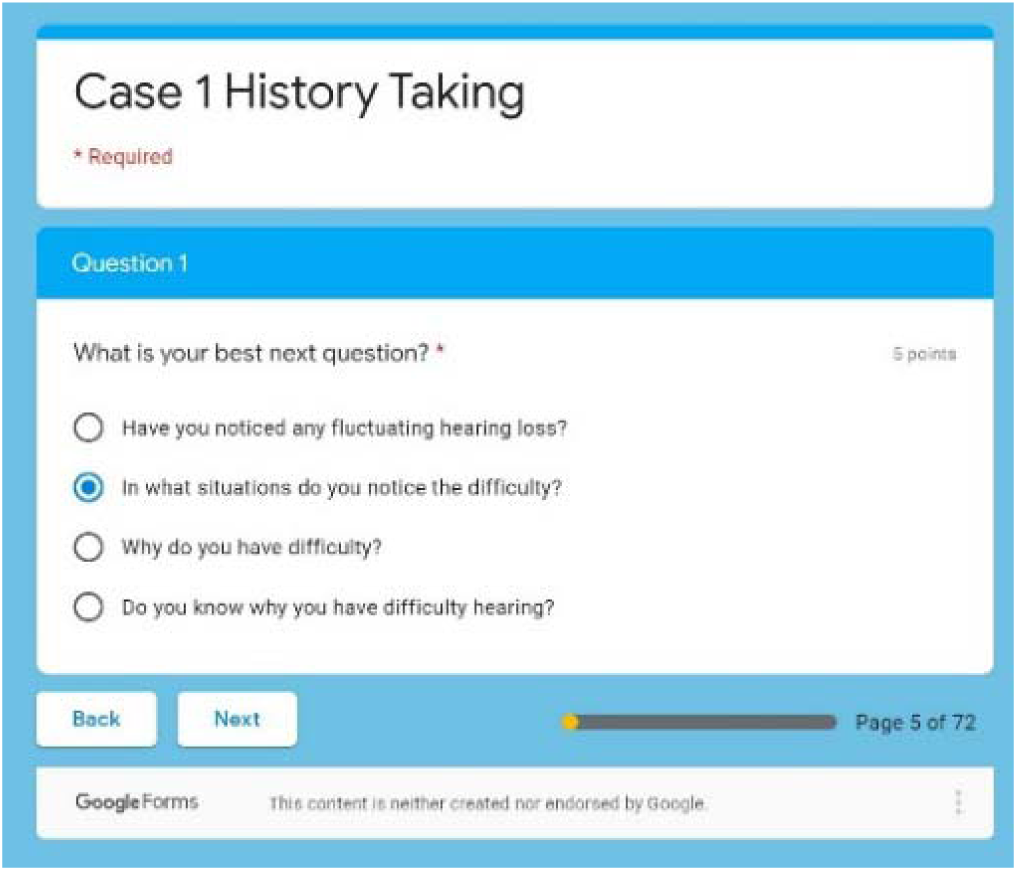
Screenshots of the history-taking session.

The second section in the simulation was about examination and investigation. In this section, the student accessed a simulation of tests and devices used in diagnosing audiology cases such as the Pure Tone Audiometry, the Tympanometry, and the Bone Conduction Tests (See Figures 8, and 9). This session was different from the previous one. Here the student uses their clinical skills. They must test the hearing acuity and identify the hearing loss. This session was also supported by hard scaffolds in the form of pre-scripted test results to allow the student to know the right examination results after they do it. However, and like the previous activity, any wrong answer was counted negatively in the overall grade. This way of grading measured the clinical reasoning skills of the student and enabled the student to reach the end of the simulation.

**Figure 4.**
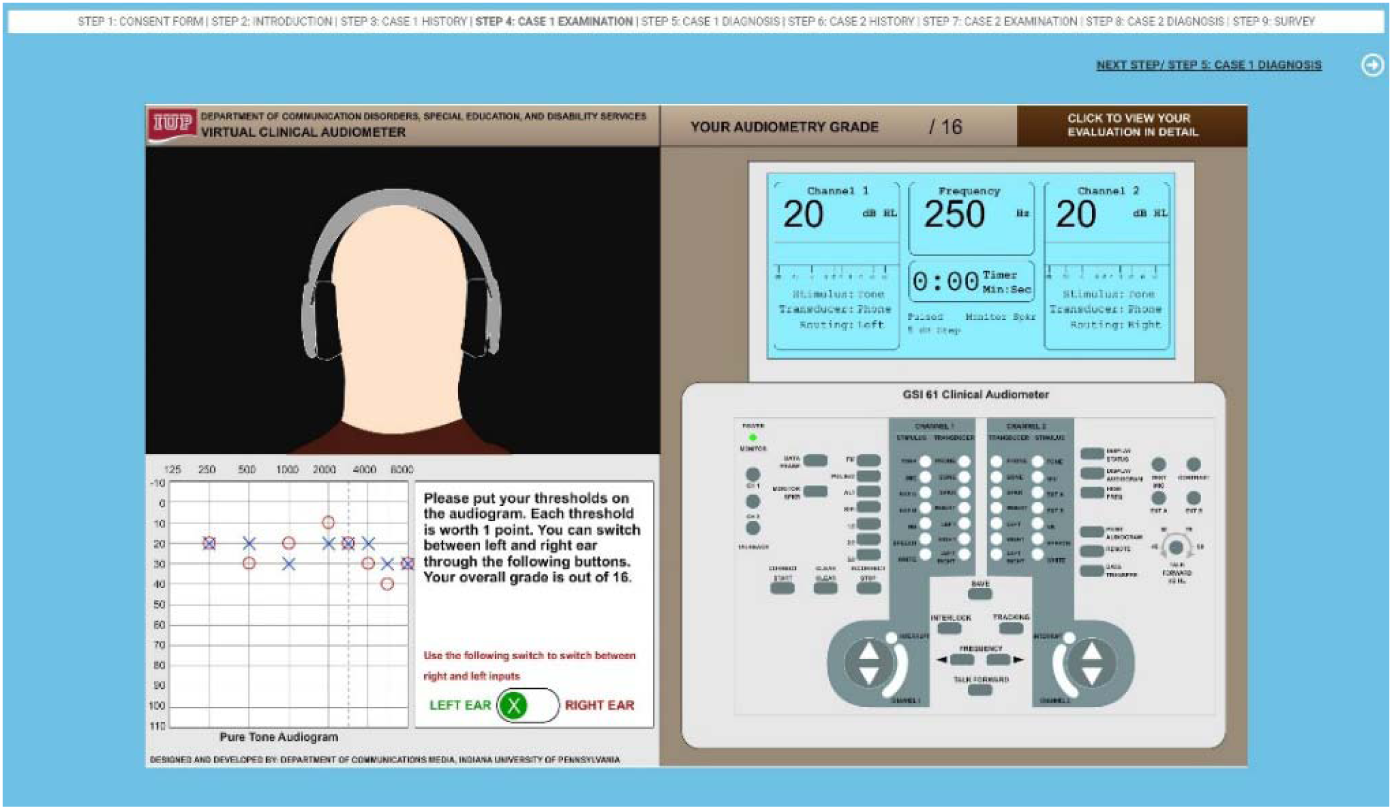
Audiogram simulation session.

**Figure 5.**
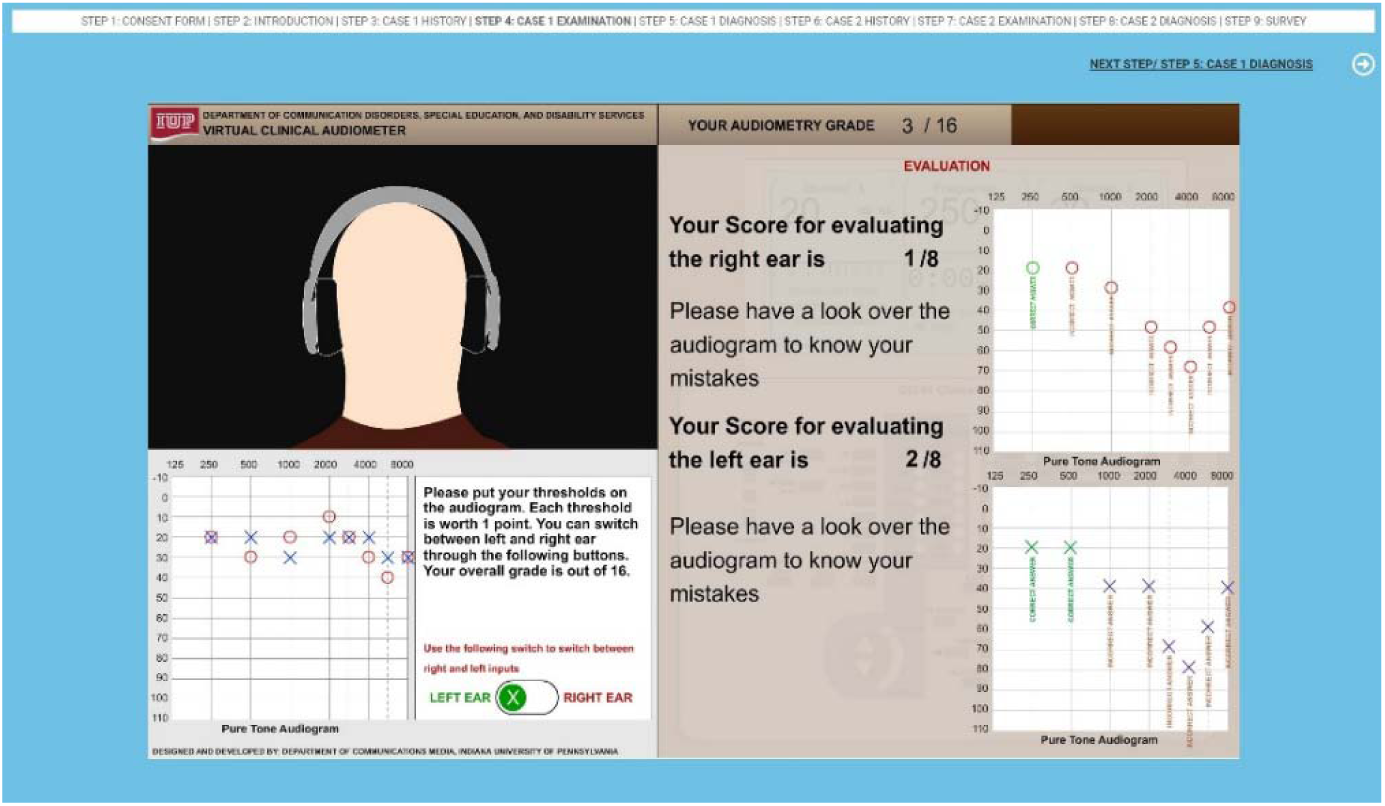
Screenshots of the examination session.

**Figure 6.**
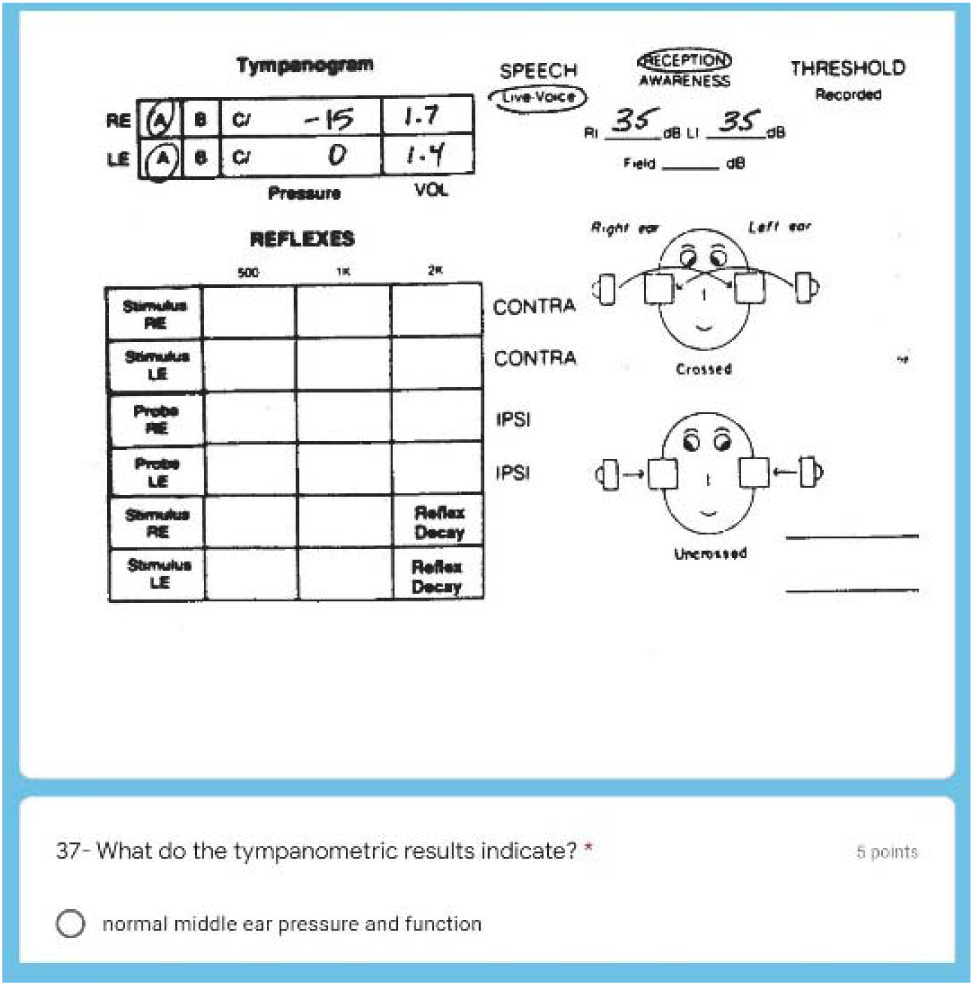
Diagnosis section - Typmanometric findings.

**Figure 7.**
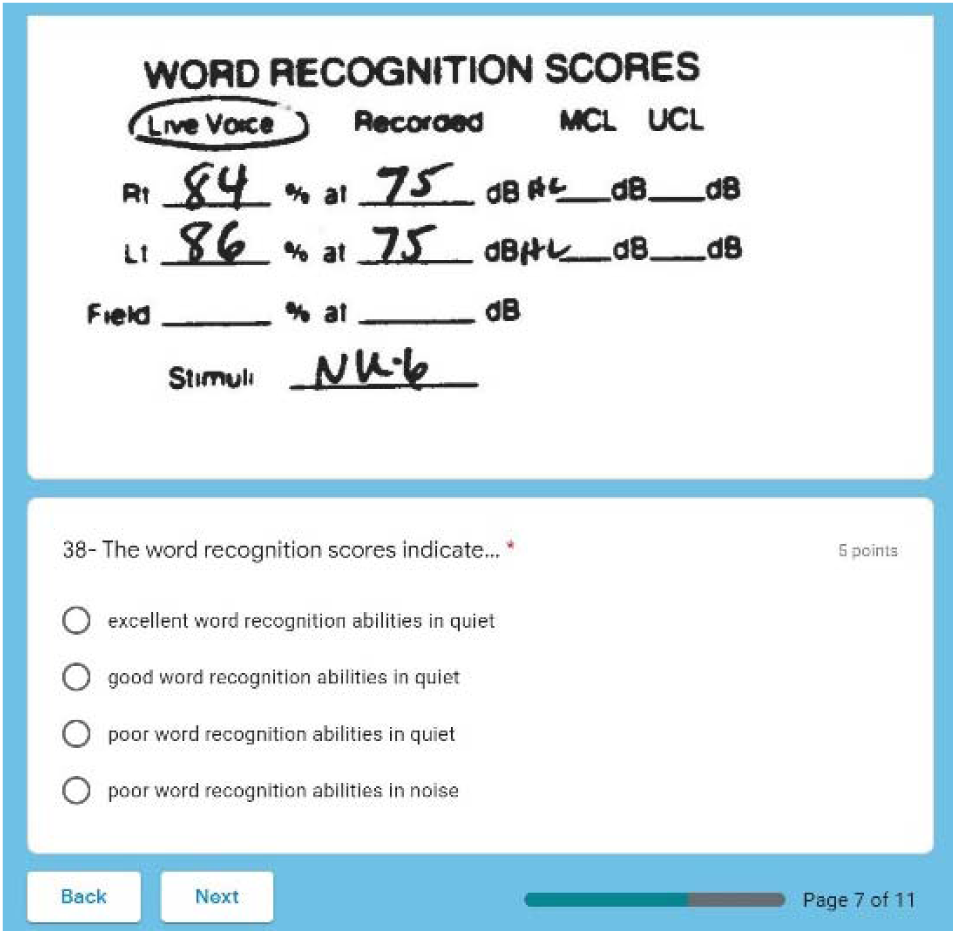
Screenshot of the diagnosis session.

**Figure 8.**
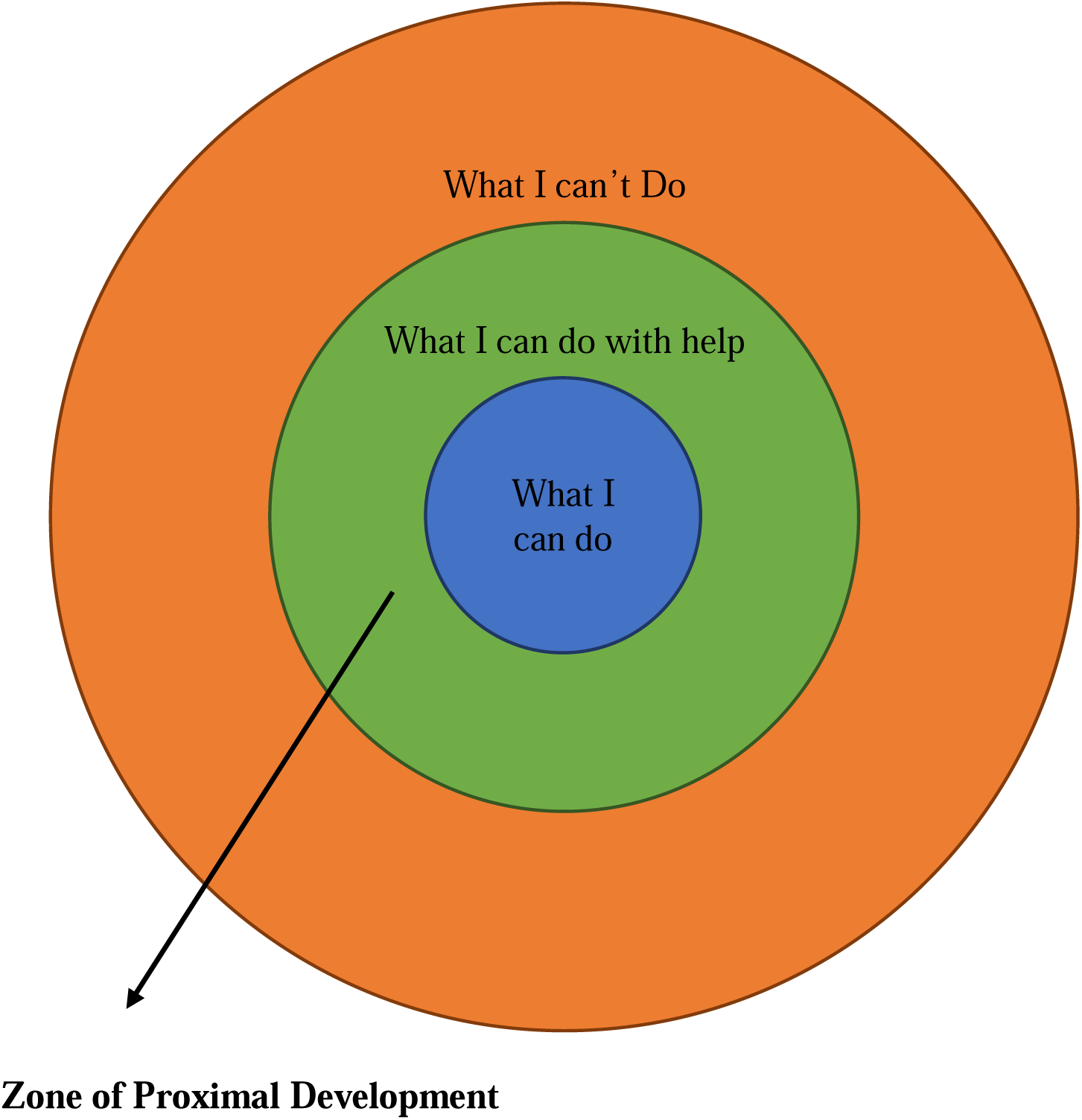
Zone of Proximal Development.

The third section was another set of problem-solving multiple-choice questions, with hard scaffolds aimed to engage the student in a clinical reasoning process to diagnose the case based on the history and the examination of the patient (See Figures 10, and 11). The final question in this session was a question about the final diagnosis of the case. Whether or not the student gets the right final diagnosis, customized feedback was given at the end with a detailed discussion about the case and how this final diagnosis was decided.

After finishing the first case, the program asked the student to proceed to the second case. The second case had the same sequence but was more challenging.

### The Experiment

During the experiment, three groups were included. The first group was the control group that was provided with virtual cases with no collaboration. Members of this group were not allowed to talk to each other while solving the cases, and students are only allowed to solve the cases individually. The second group was given the same virtual cases aided with live collaboration, which means they were allowed to solve the cases through live, in-lab discussions collaboratively. The third group was given virtual cases aided with a virtual collaboration tool in the form of a chatroom that enabled them to collaborate virtually in solving the cases. The clinical reasoning skills were determined through analyzing the scores resulting from the virtual cases.

Since there were three groups for this experiment, three versions of this simulation program were developed. The first version was the placebo one. This version was given to the control group, and it did not have any collaboration tool. The students in the control group solved the two cases individually and were not allowed to collaborate to solve the cases. Every step in the program is given grading points, and at the end of the simulation session, the points were added up, and a final score was assigned to the students in an excel sheet

The second version was the simulation program that was aided with live collaboration. In this version, students were given the same simulation version that does not have a collaboration tool but was allowed to talk to each other in the lab and collaborate in person with their classmates to solve the cases. The two cases are the same across all the versions.

The third version was the one with virtual collaboration. In this version, the same simulation program was used and virtual collaboration tools in the form of a chatroom, and file-sharing tools are embedded in the program. This simulation was assigned to the third experimental group, and students were asked to collaborate via the online collaboration tools embedded in the program. These computer simulations can grade students and provide an overall score on their performance. The grades included detailed points for each section of the simulation.

Three proctors were responsible for the three labs. Each proctor was given a paper with instructions specific to each group. The duration of the experiment was 45-60 minutes. At the beginning of the session, the proctor read the instructions loudly to the students; then, they got the consent to sign. The consent contained the primary information about the study and gave a choice to the student to withdraw without any negative consequences on their grades. The students then started doing the simulation in the three separate labs. The proctors made sure everything was clear and answered any questions students had during the experiment.

The progress of each student was recorded in the simulation program. Each section was graded separately but was linked to the student’s email address. The email address was used as an identifier, so the researcher can link all student’s grades in all sections when the data collection begins. After assigning all the grades to the student’s email address, the email address was then deleted to destroy any identifiable information about the students.

To make sure the experiment was balanced between the two universities, the clinical simulation designed had the questions embedded into it. The same two modules were given to the students of both universities. Fortunately, the same instructor designed the classes in both universities as the instructor was working on one of the universities and then got a job and moved to the second university. So, what students got during the semester was the same across both universities. With unifying the assignments across the two universities, the research then maintained the balance between both experiments.

Demographics collected during the experiment were age, and gender during the survey to get descriptive data about the subjects participating in this experiment. GPA from the instructors with permission from the students was collected to see whether or not the results are affected by this factor.

### Power Analysis

Before recruiting the subjects for this study, a power analysis was conducted to determine the appropriate sample size for the experiment. The least effect size was found to be .82 after reviewing the literature. The sample size was calculated using G*Power software taking into consideration multiple parameters; power value that equals .9, effect size that equals .82, and a confidence level that equals 95 percent. The calculated required sample size was then 24.

### Sampling and Recruitment

Undergraduate students of the Department of Communications Disorders, Special Education, and Disability Services at the Indiana University of Pennsylvania, and the Department of Communications Disorders at Wichita State University were the subjects of this study. In order to conduct this study in the two universities, the researcher went through the IRB process for both universities and got the IRB approvals needed for the study. For the Indiana University of Pennsylvania, the researcher had access to the students through the chair of the department. The chair of the department was responsible for communicating with the instructor of the audiology classes, where students are needed for the study. For Wichita State University, a faculty from the Department of Communication Disorder was responsible for communicating with the students there.

The sample was a convenience sample due to the nature of the study and the availability of audiology cases. The recruitment process started with contacting the advisors in both departments, giving them an idea about the study and asking them to help to support the study by inviting their students to the experiment. After getting permission from the instructors of the audiology classes, students were invited to participate in the study. The instructors encouraged the students to participate in the study by giving them bonus points. However, instructors were asked to provide alternative ways for those who did not participate to earn these bonus points.

Undergraduate students taking audiology classes at University One were 22. At University Two, there were 16 students taking audiology classes. The aim of this study was to examine the effect of collaboration on audiology students’ clinical reasoning skills at two universities. The reason behind conducting the experiment at two universities was to insure getting the minimum required sample size since there was one audiology class offered per semester in each university.

After inviting the students, the instructor was given the letter of participation via an online link, and students recruited were contacted with the date and the time of the experiment. The researcher reserved three labs for the three experimental groups. After gathering the participant in one lab, there were randomly assigned to one of the three groups. Each group then moved to one of the three reserved labs. The experiment in the three labs is held in parallel at the same time.

## Power Analysis

Before recruiting the subjects for this study, a power analysis was conducted to determine the appropriate sample size for the experiment. To do the power analysis, the effect size was needed. The least effect size was found to be .82 after reviewing the literature. The sample size was calculated using G*Power software taking into consideration multiple parameters; power value that equals .9, effect size that equals .82, and a confidence level that equals 95 percent. The calculated required sample size was then 24.

## Demographic Findings

The subjects for this study were undergraduate students taking audiology classes. The experiment was conducted at two locations; one at a public, medium-sized, teaching university in Western Pennsylvania (University One), and the other one at a public, medium-sized, research university in Kansas (University Two). One audiology class is typically offered at each university every semester. Subjects taking the audiology class in each university were invited to participate in the study.

The demographics collected in this study included gender, age, and the subject’s university. Out of 24 subjects taking the audiology class at University One, 22 subjects participated in the study. Out of 18 subjects taking the audiology class at University Two, 16 subjects participated in the study. The total number of subjects was 38 (See Table 2).

**Table 1.**
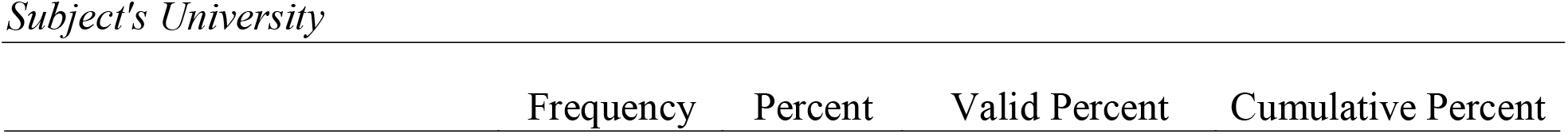
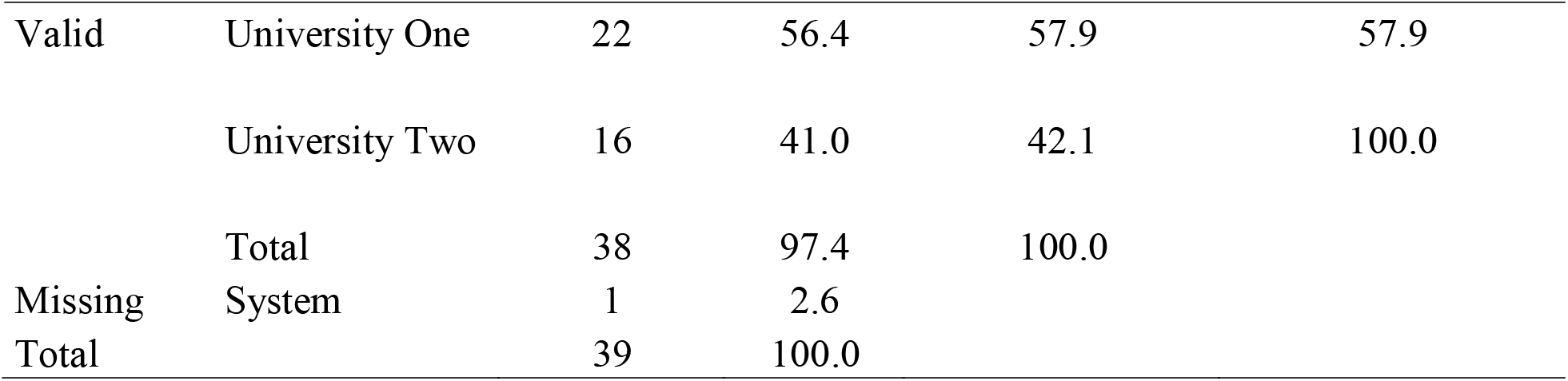

|  | Frequency | Percent | Valid Percent | Cumulative Percent |
| --- | --- | --- | --- | --- |

|  |  |  |  |  |  |
| --- | --- | --- | --- | --- | --- |
| Valid | University One | 22 | 56.4 | 57.9 | 57.9 |
|  | University Two | 16 | 41.0 | 42.1 | 100.0 |
|  | Total | 38 | 97.4 | 100.0 |  |
| Missing | System | 1 | 2.6 |  |  |
| Total |  | 39 | 100.0 |  |  |

**Table 2.**

|  |  | Frequency | Percent | Valid Percent | Cumulative Percent |
| --- | --- | --- | --- | --- | --- |
| Valid | Male | 2 | 5.1 | 5.3 | 5.3 |
|  | Female | 36 | 92.3 | 94.7 | 100.0 |
|  | Total | 38 | 97.4 | 100.0 |  |
| Missing | System | 1 | 2.6 |  |  |
| Total |  | 39 | 100.0 |  |  |

The gender information was one of the three demographic information collected from the subjects. In this study, 38 subjects participated in the study, two males and 36 females (See Table 3). While the gender representation was not balanced in the study, it was impossible to overcome this imbalance because the subjects taking audiology classes at both universities were mostly females. The age of the subjects was 84.6% from 18-24 years, and 12.8% from 25-34 years, which is logical given the fact that the subjects were undergraduate (See Table 4).

**Table 3.**
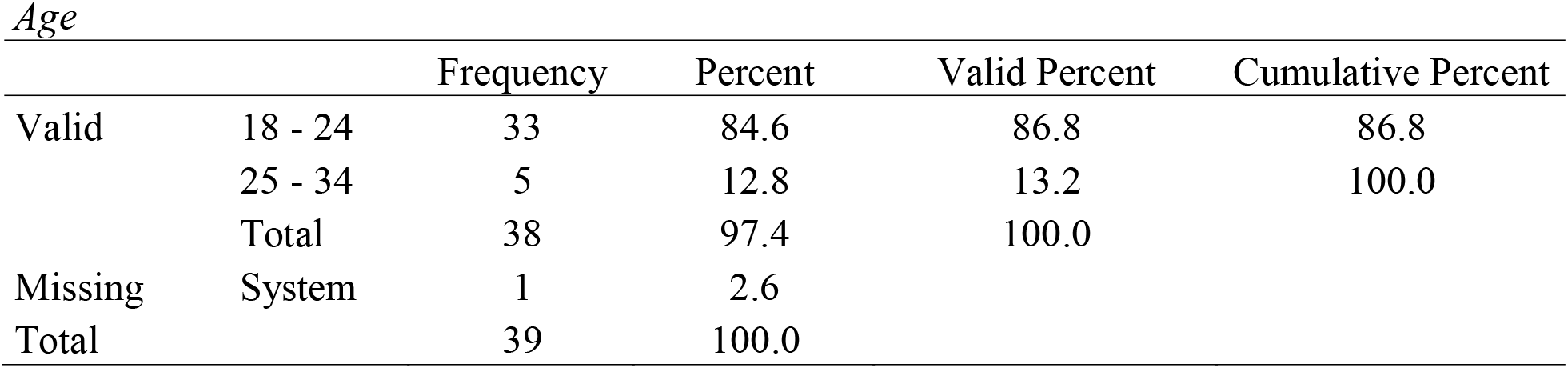

**Table 4.**

*Treatment Groups*
|  |  | Frequency | Percent | Valid Percent | Cumulative Percent |
| --- | --- | --- | --- | --- | --- |
| Valid | No Collaboration | 14 | 36.8 | 36.8 | 36.8 |
|  | Virtual Collaboration | 12 | 31.6 | 31.6 | 68.4 |
|  | Live Collaboration | 12 | 31.6 | 31.6 | 100.0 |
|  | Total | 38 | 100.0 | 100.0 |  |

## The Experiment

The subjects were divided into three treatment groups: no collaboration group, virtual collaboration group, and live collaboration group. There were 14 subjects in the no collaboration group, 12 in the virtual collaboration group, and 12 in the live collaboration group (See Table 5). These percentages show that the treatment groups were divided nearly evenly.

**Table 5.**
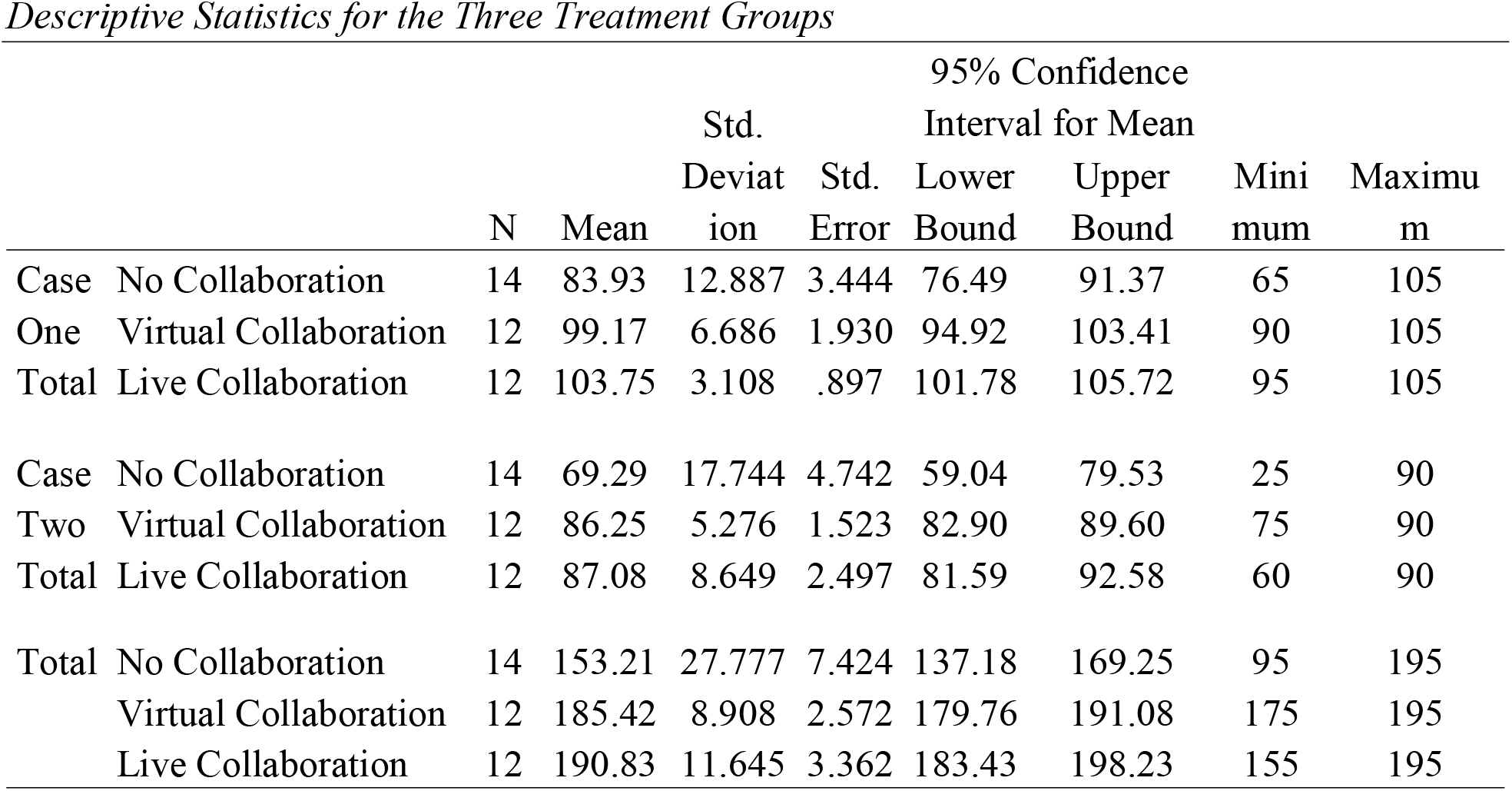

### The difference between no collaboration, virtual collaboration, and live collaboration treatments

The no collaboration group consisted of 14 subjects and had a mean score of 83.93 out of 105 (80 percent) in Case One, 69.29 out of 90 (77 percent) in Case Two, and a total of 153.21 out of 195 (78.6 percent). The virtual collaboration group consisted of 12 subjects and had a mean score of 99.17 out of 105 (94.4 percent) in Case One, 86.25 out of 90 (95.8 percent) in Case Two, and a total of 185.42 out of 195 (95.1 percent). The live collaboration group consisted of 12 subjects and had a mean score of 103.75 out of 105 (98.8 percent) in Case One, 87.08 out of 90 (96.8 percent) in Case Two, and a total of 190.83 out of 195 (97.9 percent) (See Table 6).

**Table 6.** *One-Way ANOVA for the Three Treatment Groups*

|  |  | Sum of Squares | df | Mean Square | F | Sig. |
| --- | --- | --- | --- | --- | --- | --- |
| Case One Total | Between Groups | 2843.155 | 2 | 1421.577 | 18.048 | .000 |
|  | Within Groups | 2756.845 | 35 | 78.767 |  |  |
|  | Total | 5600.000 | 37 |  |  |  |
| Case Two Total | Between Groups | 2675.345 | 2 | 1337.672 | 8.966 | .001 |
|  | Within Groups | 5222.024 | 35 | 149.201 |  |  |
|  | Total | 7897.368 | 37 |  |  |  |
| Total | Between Groups | 10952.428 | 2 | 5476.214 | 15.463 | .000 |
|  | Within Groups | 12394.940 | 35 | 354.141 |  |  |
|  | Total | 23347.368 | 37 |  |  |  |

Analysis of Variance test was used to answer the first research question. The results showed that in the first case, the F value was 18.048 and the p-value was .000 which is less than .05. With these results, the researcher was able to reject the null hypothesis. The decision is that in the first case, there is a statistically significant difference in terms of clinical reasoning skills between subjects using virtual patients aided with no collaboration, live collaboration, and virtual collaboration (df = 2, F = 18.05, P < .05) (See Table 7).

**Table 7.**
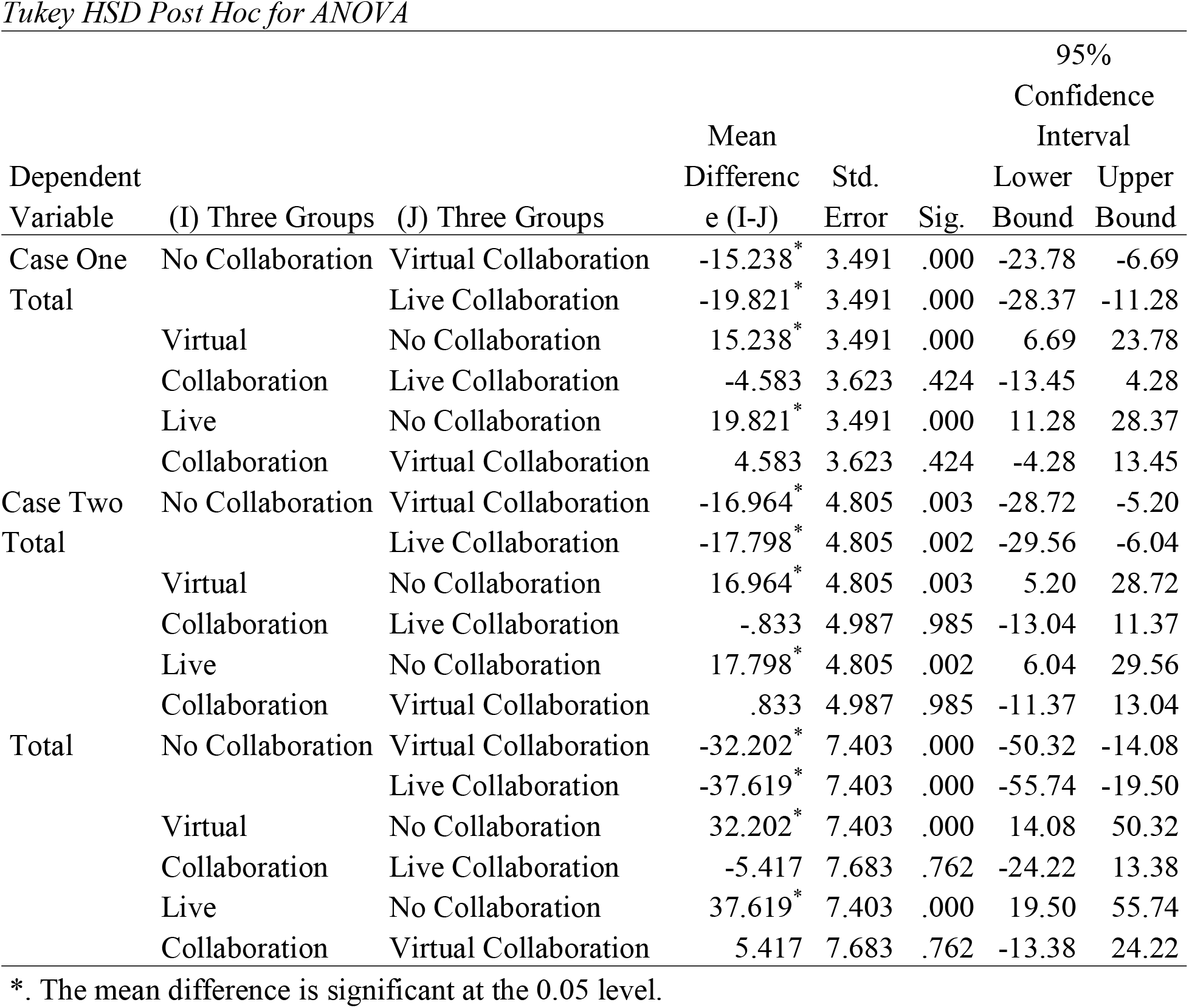

In the second case, the F value was 9.966. With these results, the researcher was able to reject the null hypothesis. The decision is that in the second case, there is a statistically significant difference in terms of clinical reasoning skills between subjects using virtual patients aided with no collaboration, live collaboration, and virtual collaboration (df = 2, F = 8.966, P < .05) (See Table 7).

When the two cases are summed up and a total score obtained, one-way ANOVA gave an F value of 15.463 and a P-value of .000. With these results, the researcher was able to reject the null hypothesis. The decision the researcher reached from the results when completing the simulation indicated a statistically significant difference in terms of clinical reasoning skills between subjects using virtual patients aided with no collaboration, live collaboration, and virtual collaboration (df = 2, F = 15.463, P < .05) (See Table 7).

The analysis of variance indicated a statistically significant difference between the three treatment groups in terms of clinical reasoning skills. To conduct a more detailed analysis, the researcher used Tukey as a post hoc test for the analysis of variance to examine the following: the difference in clinical reasoning skills between subjects used virtual patients aided with no collaboration and with collaboration, the difference in clinical reasoning skills between subjects used virtual patients aided with no collaboration and virtual collaboration, the difference in clinical reasoning skills between subjects used virtual patients aided with no collaboration and live collaboration, and the difference in clinical reasoning skills between subjects used virtual patients aided with virtual collaboration and live collaboration.

### No collaboration versus virtual collaboration treatment

For case one, the no collaboration treatment group’s clinical reasoning score was 15.238 lower than the clinical reasoning score of the virtual collaboration treatment group, and this difference was statistically significant. For case two, the no collaboration treatment group’s clinical reasoning score was 16.964 lower than the clinical reasoning score of the virtual collaboration treatment group and this difference was statistically significant (p<.05). For the overall simulation, the no collaboration treatment group’s clinical reasoning score was 32.202 lower than the clinical reasoning score of the virtual collaboration treatment group, and this difference was statistically significant (See Table 8).

### No collaboration versus live collaboration treatment

For case one, the no collaboration treatment group’s clinical reasoning score was 19.821 lower than the clinical reasoning score of the live collaboration treatment group and this difference was statistically significant. For case two, the no collaboration treatment group’s clinical reasoning score was 17.798 less than the clinical reasoning score of the live collaboration treatment group and this difference was statistically significant. For the overall simulation, the no collaboration treatment group’s clinical reasoning score was 37.619 less than the clinical reasoning score of the live collaboration treatment group and this difference was statistically significant (See Table 8).

### Virtual collaboration versus live collaboration treatment

For case one, the virtual collaboration treatment group’s clinical reasoning score was 4.583 less than the clinical reasoning score of the live collaboration treatment group but this difference was statistically not significant (p>.05). For case two, the virtual collaboration treatment group’s clinical reasoning score was .833 less than the clinical reasoning score of the live collaboration treatment group but this difference was statistically not significant (p>.05). For the overall simulation, the virtual collaboration treatment group’s clinical reasoning score was 5.417 less than the clinical reasoning score of the live collaboration treatment group but this difference was statistically not significant (p>.05) (See Table 8).

In conclusion, Tukey HSD Post Hoc test for One-Way ANOVA provided more in-depth understanding of the results. The results showed that live collaboration treatment group was got the highest scores. These scores were at a significant level compared to the no collaboration group, but they were not at a significant level compared to the virtual collaboration treatment.

## Results Discussion

The significant results obtained were expected as explained in the literature review section. Collaboration enhances clinical reasoning skills in two ways: creating a collaborative reasoning environment, and scaffolding students’ learning process (Gonulal & Loewen, 2018). Using collaborative reasoning enhances student’s decision making when dealing with a clinical case as collaborative reasoning is found to be more effective than individual reasoning during the process of diagnosing clinical cases (Dewhurst, Borgstein, Grant, & Begg, 2009). This was consistent with the study’s findings.

When looking at the mean scores of the given audiology simulation, the no collaboration group, had a mean score of 83.93 for the first virtual case, which was 15.24 points lower than the mean score of the virtual collaboration treatment group and 19.82 points lower than the live collaboration treatment group. For the second case, the mean score for the no collaboration treatment group was 69.29, which was 16.94 points lower than the virtual collaboration treatment group, and 17.8 points lower than the live collaboration treatment group. The total score for the no collaboration group was 153.21, which was 32.2 points lower than the virtual collaboration treatment group and 37.6 points lower than the live collaboration treatment group.

These differences were significant when running One-way ANOVA. The no collaboration treatment group used individual reasoning to solve the cases, while virtual and live collaboration treatment groups used collaborative reasoning to solve the cases. From these findings, it is obvious that collaborative reasoning was associated with higher performance scores, while individual reasoning was associated with lower performance scores.

### Collaboration as a Medium in the Situated Learning Realm

When running a one-way ANOVA test on the performance scores of the audiology simulation, there was a significant difference between the three treatment groups. To get more insight into the data and to see exactly which group has significant differences with other groups, the Tukey post hoc test was run. When running post hoc Tukey HSD test for ANOVA, there were significant differences between no collaboration and virtual collaboration groups, and no collaboration and live collaboration groups. However, there was no significant difference between virtual collaboration treatment and live collaboration treatment. These results can be explained through the situated learning theory.

As reviewed in chapter two, Situated Learning Theory states the acquired knowledge should be put in a specific context (Mattar, 2018). To avoid inert knowledge, which is the non-usable knowledge the individual might have, it is important to situate any acquired knowledge in a specific social context (Pérez-Sanagustín, Muñoz-Merino, Alario-Hoyos, Soldani, & Kloos, 2015). When addressing clinical reasoning, the learner uses clues to find an accurate clinical diagnosis. The information and clues gathered should be put into a reasonable context to be successfully interpreted. (Pérez-Sanagustín et al., 2015) This process, when it happens in the clinical context, is called the clinical reasoning process.

In this simulation, students were given two virtual cases, these virtual cases provided the context for the clinical reasoning process. While students were able to apply the knowledge they have on the given scenario, this scenario, in the no collaboration simulation, lacks a social context. When using virtual or live collaboration with the simulation, the clinical situation now has the social context needed. The effect was obvious when the difference became significant between each collaboration group and the no collaboration one.

To understand how collaboration looks like in the situated learning environment, it is important to discuss the term community of practice. A community of practice is a concept that appeared associated with the idea of situated learning (Cruess, Cruess, & Steinert, 2018). A community of practice is a group of people who share the same interests. The community of Practice represents the social learning spaces in the situated learning environment (Nicolini, Scarbrough, & Gracheva, 2016). When it comes to social learning, social interaction, and collaboration become the medium for this process (Mattar, 2018). This study used collaboration as a method to create virtual as well as physical communities of practices, which are found to be crucial in the healthcare practice (Nicolini et al., 2016). In this study, audiology students were asked to act as communities of practice. One of these communities was face-to-face and the other was virtual. The above two concepts, situated learning, and community of practice go along with the significant results obtained in this study.

### Collaboration as a Fuel to the Social Development

During the simulation experience, two treatment groups were asked to engage in collaborative reasoning experience through virtual or live social interaction. As mentioned previously, the main total score of the no collaboration treatment group was 153.21 points, which was significantly less than both the virtual collaboration group (185.42 points) and the live collaboration group (190.83 points). The only difference between the no collaboration simulation and the virtual/live collaboration simulation was social interaction. This explains that social interaction had a significant effect on audiology students’ performance. While the main live collaboration score seems to be slightly higher than the main virtual collaboration score, there was no significant difference between the two treatments. This explains that social interaction itself has a significant effect on performance no matter how this social interaction happens (live or virtual). This effect can be explained through the Social Development Theory.

Social Development Theory argues that social interaction comes before cognitive development (Vygotsky, 1980). Vygotsky’s theory was one of the foundations of constructivism (Mattar, 2018). According to the theory, the individual learns first through social interaction, and cognitive development happens first on the social level (Vygotsky, 1980). Later, cognitive development happens on the individual level. When Vygotsky (1980) introduced social development theory, he described three major concepts: role of social interaction in cognitive development, the more knowledgeable other (MKO), and the zone of proximal development (Kalina & Powell, 2009). The three concepts can help explain the process of developing clinical reasoning competency among medical learners.

Vygotsky argued that learners need to first gain knowledge through social interaction as this interaction is essential for cognitive development (Sandars, Kokotailo, & Singh, 2012). From the clinical perspective, the medical learner first needs to learn how to collect and interpret the clues to reach an accurate diagnosis (Boshuizen et al., 2018). This learning process could be enhanced by putting the learner in a social learning environment, where the learner engages in a collaborative reasoning process before developing individual competency (Sandars et al., 2012).

When looking at the current virtual patient designs, most virtual patients provide an individual learning experience to the students (Braun et al., 2019). This individual approach in developing clinical reasoning competency opposes the idea of social development. To apply social development theory in the context of clinical reasoning, it is important to engage learners in a social learning experience first, in which they develop their clinical reasoning competency (Sandars, Kokotailo, & Singh, 2012). When they become competent, they become able to develop their clinical interpreting skills individually (Sandars et al., 2012).

The second concept Vygotsky introduced is the “More Knowledgeable Other” (Vygotsky, 1980). This concept provides an argument that the learner needs someone more knowledgeable to help during the process of cognitive development (Kalina & Powell, 2009). While the original concept refers to those who are experts, or older adults, recently, this concept refers also to computers, and software that can help learners during the cognitive development process (Putman, 2017). In this study, virtual patients can be considered as MKOs as they have embedded pre-scripted feedbacks that assist students throughout the simulation experience.

The third concept in Vygotsky’s theory is the Zone of Proximal Development (ZPD) (Vygotsky, 1980). ZPD refers to the area between what is known and what is unknown (Figure 9). Vygotsky argues that learning occurs in the zone of proximal development area. In this zone, the learner needs help to pass it (Chaiklin, 2003). In this study, scaffolds provided the support needed in this stage. This concept explains how subjects of this study had significantly higher scores when using collaboration as an assistive technique during the simulation experience.

### Collaboration as a Ladder in the Scaffolding Process

When looking at the difference between the three treatment groups, it is obvious that two groups have collaboration and one group that has no collaboration. As explained in the literature, collaboration is considered a soft scaffold (Braun et al., 2019). The soft scaffold is dynamic, customizable, and changeable according to the user’s input (Gonulal & Loewen, 2018).

Collaboration is a good application of soft scaffolding, where peers act as units in this scaffolding process (Lu, Lajoie, & Wiseman, 2010). ANOVA, as well as Tukey results in this study were significant, which supports the assumption based on the literature review, which explains how collaboration was effective in enhancing student’s performance. While this effect may be explained through theories of situated learning and social development, it also can be examined through scaffolding and collaborative learning.

Looking at the collaboration from a different perspective, it is considered one of the useful scaffolding tools (Braun et al., 2019). Scaffolding means assisting the learner through the learning process until reaching the desired learning outcome (Shin, Brush, & Glazewski, 2017). There are two forms of scaffolding: hard scaffolds, which are the fixed pre-scripted scaffolds; and soft scaffolds, which are the interactive, customizable ones (Shin, Brush, & Glazewski, 2017). In this situation, collaboration is considered a soft scaffold as collaboration is not a fixed process and is customizable according to the context (Gonulal & Loewen, 2018).

Considering the clinical reasoning process is an organized step-by-step process and not a holistic one, scaffolding can be used effectively to support this process (Braun et al., 2019). That is what happened in this experiment. During the simulation experience, the two treatment groups were put into a collaborative learning environment and asked to use this treatment to solve each step of the virtual cases to make sure the collaboration is used as a scaffolding approach.

Collaborative Learning is an educational approach that involves groups of students working together to solve a problem (Lin, Hou, & Chang, 2020). From a technological perspective, collaborative learning is a common approach in computer-based learning. The concept of Computer-Supported Collaborative Learning (CSCL) is a field of technology based on the concept of collaborative learning and is considered an application of it in the field of computer technology (Halavais, 2016). The concept has nourished after the evolvement of the student-centered approach to learning (Lu et al., 2010). Individualization was the main principle for computer-supported education in the past (Lu, Lajoie, & Wiseman, 2010). CSCL played a major role in changing the main principle of computer-supported learning from individualization to interaction (Halavais, 2016).

In this study, the collaborative learning environment can be better described as a collaborative reasoning environment. Two groups were given two different collaboration treatments: virtual collaboration, and live collaboration. Using analysis of variance, it is found that collaboration, whether virtual or live, has a positive effect on students’ clinical reasoning.

## Implications

This study implies collaboration, whether virtual or live, is effective for students’ clinical reasoning skills. It also implies that collaboration in its two forms used in this study shows no significantly different effect based on the way the collaboration is provided. The main total score for the virtual collaboration treatment group was 185.42 points (Table 6), which is 5.4 points less than the live collaboration group (190.83 points). This difference, when running Tukey post hoc test, for ANOVA was not significant.

This implication is explained using the theoretical framework used in the second chapter. Collaboration creates an active environment where students interact to perform group reasoning (Custers, 2018). This collaborative reasoning as mentioned in the second chapter reduces the intrinsic cognitive load, which in turn, enhances the learning experience (Norman et al., 2017). Collaborative reasoning is also found to be more effective than individual reasoning then dealing with clinical cases (Bateman, Allen Maggie, Kidd, Parsons, & Davies, 2012).

Scaffolding, on the other hand, explains how social interaction can affect the learning outcome. Xun and Land (2004) proposed a conceptual framework to scaffold problem-solving activities using prompts and peer interactions, which is a similar framework to the design of the simulation experience used in this study. (Shin et al., 2017) divided scaffolds into two forms: soft scaffolds and hard scaffolds.

The simulation used in this study had both soft and hard scaffolds. While there was no way to determine the effect of each scaffolding technique separately, the results of this study indicate that adding soft scaffold to the simulation experience using peer collaboration is significantly more effective in enhancing students’ clinical reasoning skills. Tukey HSD post hoc test (see table 8) highlighted the statistically significant difference between the no and virtual/live collaboration, and the insignificant difference between virtual and live collaboration. The insignificant results between the virtual and the live collaboration treatment groups give an indication for instructional designers that using virtual collaboration as a replacement for the live collaboration may not affect the overall learning experience.

## Summary

This study addressed an issue in virtual patients’ design, which is the lack of collaborative reasoning (Braun et al., 2019). Collaborative reasoning is an important part of the clinical reasoning process, which is an integral part of the medical practice. Without enhancing clinical reasoning skills, clinicians tend to commit what is called clinical error, which means reaching an inaccurate diagnosis (Norman & Eva, 2010).

So, it was important to examine how embedding collaborative approaches in the simulation experience could affect students’ performance. In this study, the target was to look at students who take audiology classes. The reason why audiology students were the target subjects for this study is twofold. First, the audiology departments were accessible for the study; second, the audiology field is one of the basic clinical fields that can provide an insight into the effect of collaboration on clinical reasoning, thus giving an example for other clinical fields. It also encourages conducting more research to examine the effect of collaboration on clinical reasoning skills in other fields.

From the above issue and interest, the purpose of this study is to examine whether there is a statistically significant difference in clinical reasoning between audiology students using virtual patients aided with no collaboration, virtual collaboration, and live collaboration. Two research questions were formed to address the effect of the collaboration treatment on both clinical reasoning. Two null hypotheses were stated for the two research questions. The study aimed to reject or to fail to reject these null hypotheses.

A quasi-experimental design was used to address the study’s purpose. The post-test only experiment was designed as a part of the study’s methodology. The subjects were divided into three treatment groups: no collaboration, virtual collaboration, and live collaboration. The simulation experience was in the form of two virtual cases to test the clinical reasoning of the students.

Two sets of data were collected, performance/clinical reasoning data. Both were analyzed using One-Way ANOVA to answer the two research questions. There were statistically significant differences in clinical reasoning skills of students taking audiology simulation aided with no collaboration, virtual collaboration, and live collaboration. Using the Tukey post hoc test for the One-Way ANOVA test, the results were significant for both virtual and live collaboration treatments. However, the result was not significant between the live and the virtual collaboration treatments.

These findings were explained through the theoretical framework used in this study. Situated learning and social development theories explain how social interaction plays an important role in cognitive development. Cognitive development happens first through social interaction (Vygotsky, 1980). The individual, after the cognitive development by social interaction, becomes competent to develop their personal interpretation for the acquired knowledge (Vygotsky, 1980). Collaborative learning and scaffolding help providing the student continuous support throughout the learning process (Shin et al., 2017).

Cognitive Load Theory explains how cognitive load can be divided into three categories: intrinsic load, extraneous load, and germane load (Van Merriënboer & Sweller, 2010). The intrinsic load can be negatively affected by the increased interactivity during the clinical learning process (Van Merriënboer & Sweller, 2010). Collaborative reasoning plays an effective role in reducing the intrinsic load, thus enhancing the learning experience (Sandars et al., 2012).

The theoretical framework used along with the results of this study suggests that collaboration, whether virtual or live can enhance students’ clinical reasoning skills. This study also provided some highlights on possible future research, which is discussed in the following section.

## Future Research and Significance of the Study

The findings in this study put the light on multiple potentially hot areas that can be topics for future research. As mentioned before, there was no significant difference between virtual collaboration and live collaboration. While this is a good indication that virtual collaboration could replace live collaboration, future studies can be conducted to propose new design concepts that could make virtual collaboration more effective than live collaboration.

Future research can be conducted to find ways to enhance clinical collaboration to give significant results. According to this study, collaboration along with scaffolding was effective on both students’ clinical reasoning.This finding could lead to more research on collaborative scaffolding.

In general, it is important to address virtual patients from the instructional design perspective and find new concepts to design virtual patients. While virtual patients are important elements in medical education, global crises such as the current COVID-19 pandemics requires researchers and designers to come up with solutions to overcome the disruption that could result from the applied strict measures in response to such crises. Social distancing has a negative effect on the current education process. Thus, improving the design of the current technology-based applications to provide a replacement for the face-to-face setting is an important step in overcoming such a difficult situation.

Since this study only addressed audiology as one of clinical specialties. It is also important to have future studies on how collaboration affect clinical reasoning of students in other clinical specialties. Another area that could be addressed is examining other forms of virtual patients. On the other hand, this study addressed clinical reasoning skills based on given test results. So, future research could address how would collaboration help in assessing a patient based on observed rather than measured symptoms.

Another point, future research can address, is the idea using individual reasoning versus collaborative reasoning among practicing professionals. Is it still better train students on collaborative reasoning while they end up practicing alone?

The last point addresses the gender representation in the study. In this study, due to the fact that audiologists have more female representation, it wasn’t possible to examine the difference in clinical reasoning based on gender. However, future research could address this topic to test with gender can be a factor that affect collaborative reasoning.

## Data Availability

The data is available upon request. To obtain a copy, please contact the author at

## References

Anderson, J. R., Reder, L. M., & Simon, H. A. (1996). Situated learning and education. Educational researcher, 25(4), 5–11. doi:10.3102/0013189X025004005

Andrade, A., Jaques, P., Vicari, R., Bordini, R., & Jung, J. (2001). A computational model of distance learning based on Vygotsky’s socio-cultural approach. Paper presented at the Proceedings of the MABLE Workshop. doi:10.1.1.118.519

Barrows, H. S., & Feltovitch, P. J. (1987). The clinical reasoning process. Medical Education, 21(2), 86–91. doi:10.1111/j.1365-2923.1987.tb00671.x

Bateman, J., Allen Maggie, E., Kidd, J., Parsons, N., & Davies, D. (2012). Virtual patients design and its effect on clinical reasoning and student experience: a protocol for a randomised factorial multi-centre study. BMC Medical Education, 12(1), 62. doi:10.1186/1472-6920-12-62

Benson, B. K. (1997). Coming to terms: Scaffolding. The English Journal, 86(7), 126–127. doi:10.2307/819879

Braun, L. T., Zwaan, L., Kiesewetter, J., Fischer, M. R., & Schmidmaier, R. (2017). Diagnostic errors by medical students: results of a prospective qualitative study. BMC Medical Education, 17(1), 191. doi:10.1542/peds.2016-1052

Boshuizen, H. P., Schmidt, H., Higgs, J., Jensen, G., Loftus, S., & Christensen, N. (2018). The development of clinical reasoning expertise. Clinical reasoning in the health professions(4th), 57–65.

Cai, Z., Eagan, B., Dowell, N., Pennebaker, J., Shaffer, D., & Graesser, A. (2017). Epistemic network analysis and topic modeling for chat data from collaborative learning environment. Paper presented at the Proceedings of the 10th international conference on educational data mining. Retrieved from: https://par.nsf.gov/servlets/purl/10060306

Chandler, P., & Sweller, J. (1991). Cognitive load theory and the format of instruction. Cognition and instruction, 8(4), 293–332. doi:10.1207/s1532690xci0804_2

Chang, C.-J., Chang, M.-H., Chiu, B.-C., Liu, C.-C., Chiang, S.-H. F., Wen, C.-T., … Lai, C.-H. (2017). An analysis of student collaborative problem solving activities mediated by collaborative simulations. Computers & Education, 114, 222–235. doi:10.1016/j.compedu.2017.07.008.

Colbert, J., & Chokshi, D. (2014). Technology in Medical Education—Osler Meets Watson. Journal of General Internal Medicine, 29(12), 1584–1585. doi:10.1007/s11606-014-2975-x

Cook, D. A., & Triola, M. M. (2009). Virtual patients: a critical literature review and proposed next steps. Medical Education, 43(4), 303–311. doi:10.1111/j.1365-2923.2008.03286.x

Cruess, R. L., Cruess, S. R., & Steinert, Y. (2018). Medicine as a community of practice: Implications for medical education. Academic Medicine, 93(2), 185–191. doi:10.1097/ACM.0000000000001826

Custers, E. J. (2018). Training clinical reasoning: Historical and theoretical background. In Principles and Practice of Case-based Clinical Reasoning Education (pp. 21–33): Springer, Cham. doi:10.1007/978-3-319-64828-6_2

Ellaway, R., Candler, C., Greene, P., & Smothers, V. (2006). An architectural model for MedBiquitous virtual patients. Baltimore, MD: MedBiquitous, 6. Retrieved from: http://groups.medbiq.org/medbiq/download/attachments/295/MVP_WhitePaper_11Sep2006.pdf

Enkenberg, J. (2001). Instructional design and emerging teaching models in higher education. Computers in Human Behavior, 17(i5-6), 495–506. doi:10.1016/S0747-5632(01)00021-8

Ertmer, P. A., & Newby, T. J. (2013). Behaviorism, cognitivism, constructivism: Comparing critical features from an instructional design perspective. Performance improvement quarterly, 26(2), 43–71. doi:10.1111/j.1937-8327.1993.tb00605.x

Geil, D. M. M. (1998). Collaborative reasoning: Evidence for collective rationality. Thinking & Reasoning, 4(3), 231–248. doi:10.1080/135467898394148

Gonulal, T., & Loewen, S. (2018). Scaffolding technique. The TESOL Encyclopedia of English Language Teaching, 1–5. doi:10.1002/9781118784235.eelt0180

Graber, M. (2005). Diagnostic errors in medicine: a case of neglect. The Joint Commission Journal on Quality and Patient Safety, 31(2), 106–113. doi:10.1016/S1553-7250(05)31015-4

Greenwald, S., Kulik, A., Kunert, A., Beck, S., Frohlich, B., Cobb, S., … Newbutt, N. (2017). Technology and applications for collaborative learning in virtual reality. Retrieved from: https://uwe-repository.worktribe.com/output/886338

Halavais, A. (2016). ComputerJSupported Collaborative Learning. The International Encyclopedia of Communication Theory and Philosophy, 1–5. doi:10.1002/9781118766804.wbiect195

Jaramillo, J. A. (1996). Vygotsky’s sociocultural theory and contributions to the development of constructivist curricula. Education, 117(1), 133–141. Retrieved from: http://search.ebscohost.com.dist.lib.usu.edu/login.aspx?direct=true&db=asn&AN=9611212689&site=ehost-live

Johnson, R. T., & Johnson, D. W. (2008). Active learning: Cooperation in the classroom. The annual report of educational psychology in Japan, 47, 29–30. doi:10.5926/arepj1962.47.0_29

Jonassen, D., Davidson, M., Collins, M., Campbell, J., & Haag, B. B. (1995). Constructivism and computer□mediated communication in distance education. American journal of distance education, 9(2), 7–26. doi:10.1080/08923649509526885

Kahneman, D. (1963). The semantic differential and the structure of inferences among attributes. The American journal of psychology, 76(4), 554–567. doi:10.2307/1419705

Kim, G. J. (2015). Human-computer interaction: fundamentals and practice. CRC press.

Kirschner, P. A., Sweller, J., Kirschner, F., & Zambrano R.J. (2018). From Cognitive Load Theory to Collaborative Cognitive Load Theory. International Journal of Computer-Supported Collaborative Learning, 13(2), 213–233. doi:10.1007/s11412-018-9277-y

Lave, J. (2009). The practice of learning. Contemporary theories of learning, 200–208.

Lave, J., & Wenger, E. (1991). Situated learning: Legitimate peripheral participation: Cambridge university press.

Leng, B. D., & Gijlers, H. (2015). Collaborative diagramming during problem based learning in medical education: Do computerized diagrams support basic science knowledge construction? Medical Teacher, 37(5), 450–456. doi:10.3109/0142159X.2014.956053

Lin Hou, H.-T., & Chang, K.-E. (2020). The development of a collaborative problem solving environment that integrates a scaffolding mind tool and simulation-based learning: an analysis of learners’ performance and their cognitive process in discussion. Interactive Learning Environments, 1–18. doi:10.1080/10494820.2020.1719163

Manning, B. H., & Payne, B. D. (1993). A Vygotskian-based theory of teacher cognition: Toward the acquisition of mental reflection and self-regulation. Teaching and Teacher Education, 9(4), 361–371. doi:10.1016/0742-051X(93)90003-Y

Mattar, J. (2018). Constructivism and connectivism in education technology: Active, situated, authentic, experiential, and anchored learning. RIED. Revista Iberoamericana de Educación a Distancia, 21(2). doi:10.5944/ried.21.2.20055

McGuire, C. H., & Babbott, D. (1967). Simulation technique in the measurement of problem-solving skills. Journal of Educational Measurement, 4(1), 1–10. doi:10.1111/j.1745-3984.1967.tb00562.x

Norman, G. R., & Eva, K. W. (2010). Diagnostic error and clinical reasoning. Medical Education, 44(1), 94–100. doi:10.1111/j.1365-2923.2009.03507.x

O’Donnell, A. M., & O’Kelly, J. (1994). Learning from peers: Beyond the rhetoric of positive results. Educational Psychology Review, 6(4), 321–349. doi:10.1007/BF02213419

Pelaccia, T., Tardif, J., Triby, E., & Charlin, B. (2011). An analysis of clinical reasoning through a recent and comprehensive approach: the dual-process theory. Medical education online, 16, 10.3402/meo.v3416i3400.5890. doi:10.3402/meo.v16i0.5890

Pérez-Sanagustín, M., Muñoz-Merino, P. J., Alario-Hoyos, C., Soldani, X., & Kloos, C. D. (2015). Lessons learned from the design of situated learning environments to support collaborative knowledge construction. Computers & Education, 87, 70–82. doi:10.1016/j.compedu.2015.03.019

Rencic, J., Trowbridge, R., Fagan, M., Szauter, K., & Durning, S. (2017). Clinical reasoning education at US medical schools: Results from a National Survey of Internal Medicine Sclerkship directors. Journal of General Internal Medicine, 32(11), 1242–1246. doi:10.1007/s11606-017-4159-y

Shin, S., Brush, T. A., & Glazewski, K. D. (2017). Designing and implementing web-based scaffolding tools for technology-enhanced socioscientific inquiry. Journal of Educational Technology & Society, 20(1), 1–12. Retrieved from: https://www.jstor.org/stable/pdf/jeductechsoci.20.1.1.pdf?casa_token=gmD-kep0oPIAAAAA:D5gAIcrtNnSenkaQZ-150dtMX4GxUJuHFtoMhvvXeHC2nWce3f07b1iK_ojPXCrunahBcveWTI9z9MjSiTNlcR0nueSO4XSSH2K-FrZJMZXjaY35CnK9

Simmons, B. (2010). Clinical reasoning: concept analysis. Journal of Advanced Nursing, 66(5), 1151–1158. doi:10.1111/j.1365-2648.2010.05262.x

Somerville, M., & Green, M. (2011). A pedagogy of “organized chaos”: Ecological learning in primary schools. Children Youth and Environments, 21(1), 14–34. Retrieved from: http://www.jstor.org/stable/10.7721/chilyoutenvi.21.1.0014

Stahl, G. (2005). Group cognition in computerJassisted collaborative learning. Journal of Computer Assisted Learning, 21(2), 79–90. doi:10.1111/j.1365-2729.2005.00115.x

Streveler, R. A., & Menekse, M. (2017). Taking a closer look at active learning. Journal of Engineering Education, 106(2), 186–190. doi:10.1002/jee.20160

Sweller, J. (2011). Chapter two-Cognitive Load Theory. In J. P. Mestre & B. H. Ross (Eds.), Psychology of Learning and Motivation (Vol. 55, pp. 37–76): Academic Press. doi:10.1016/B978-0-12-387691-1.00002-8

Vygotsky, L. S. (1980). Mind in society: The development of higher psychological processes: Harvard university press.

Wason, P. C., & Evans, J. S. B. (1974). Dual processes in reasoning? Cognition, 3(2), 141–154. Retrieved from: http://citeseerx.ist.psu.edu/viewdoc/download?doi=10.1.1.640.5879&rep=rep1&type=pdf

Wegner, D. M., Giuliano, T., & Hertel, P. T. (1985). Cognitive interdependence in close relationships. In Compatible and incompatible relationships (pp. 253–276): Springer.

Whitehead, A. N. (1929). The function of reason. doi:10.1007/978-1-4612-5044-9_12

Young. (1995). Assessment of situated learning using computer environments. Journal of Science Education and Technology, 4(1), 89–96. doi:10.1007/BF02211586

